# Surveillance-to-Diagnostic Testing Program for Asymptomatic SARS-CoV-2 Infections on a Large, Urban Campus - Georgia Institute of Technology, Fall 2020

**DOI:** 10.1101/2021.01.28.21250700

**Authors:** Greg Gibson, Joshua S. Weitz, Michael P. Shannon, Benjamin Holton, Anton Bryksin, Brian Liu, Sandra Bramblett, JulieAnne Williamson, Michael Farrell, Alexander Ortiz, Chaouki T. Abdallah, Andrés J. García

## Abstract

A SARS-CoV-2 testing program combining pooled saliva sample surveillance leading to diagnosis and intervention surveyed over 112,000 samples from 18,029 students, staff and faculty, as part of integrative efforts to mitigate transmission at the Georgia Institute of Technology in Fall 2020. Cumulatively, 1,508 individuals were confirmed diagnostically. The surveillance strategy, including focused intensification of testing given case clusters, was effective in disrupting transmission following rapid case increases upon entry in August 2020, and again in November 2020. Owing to broad adoption by the campus community, the program protected higher risk staff while allowing some normalization of research activities.

## Background

In mid-July 2020, the Georgia Institute of Technology committed to a comprehensive viral surveillance program aiming to mitigate the spread of SARS-CoV-2 among the on-campus student population, as well as staff and faculty. The program included the capacity to test over 1,500 individuals per day initially, rising to a maximum of 2850 per day. With an estimated 7,370 students in residence (approximately 1,170 in 38 Greek houses and 6,200 in 48 Residence Halls), and up to 5,000 staff and non-resident students visiting campus daily, each individual could be tested at least every 10 weekdays, namely biweekly. The import of ∼50-100 cases upon start of the Fall semester was anticipated in light of estimated prevalence amidst a late summer surge in the Southeast (*1*). Identifying and restricting the spread of cases in a high-density live-learn environment formed the basis for the design and scope of the surveillance program.

Assuming an effective reproduction number, R_eff_, of ∼1.5 and a characteristic growth rate of approximately one week, then 100 imported cases in the first week could lead to 150 new cases in the second week, 225 in the third week, and so on. Hence, in this scenario the surveillance program would need to identify and isolate ∼50 cases by screening 5,000 students per week using a sensitive test and assuming 1% incidence, with no more than 48-hour turnaround (and ideally next-day turnaround) resulting in isolation of positives for 10 days per CDC guidelines (*2*). University System of Georgia governance did not allow mandatory testing, but it was nevertheless projected that 70% participation biweekly would reach our goal and help us to maintain a steady-state close to 1% incidence across the semester.

The scope of the program can be illustrated by evaluating the synergistic effectiveness of non-pharmaceutical interventions (NPIs) and testing (*3,4*). NPIs reduce baseline expectations for R_eff_ such that the benefits of testing increase with reduced spread. Since the average time between infection and recovery (including both latent and infectious period) is estimated to be approximately 7-10 days (*5*), rapid testing and isolation that occurs at a similar timescale can constitute a form of mitigation. Identification and isolation of individuals via surveillance testing reduces the number of potential transmission events to susceptible individuals. Our scenario-based epidemic model analysis, consistent with other reports (*6*), indicated that achieving testing even at 10-day intervals combined with mask wearing, online courses, and distancing, could restrict the attack rate to ∼1,000 cases as opposed to >5,000 in the absence of testing.

## Methods

For safety and simplicity, a saliva-based test was developed, and used with informed consent of all participants. Individuals spit into a plastic cup, transfer 0.4 mL via a dropper into a 1.5mL Sarstedt screw-top tube, containing 0.4mL viral deactivation buffer, while preserving viral RNA for subsequent analysis. The testing procedure is supervised but self-administered and takes approximately 2 minutes to complete. In contrast, a nasal swab test was expected to be less acceptable to most students, would require storage in live viral transport medium, and involve more labor-intensive collection and laboratory processing. Two members of the team (MF and MS) had been engaged with the Georgia State testing taskforce for several months and were able to conduct a field study south of Atlanta in July, in conjunction with partners at other USG universities. The saliva-based test prototype had >95% sensitivity and specificity that met the requirements for an Emergency Use Authorization submission to the FDA. Approval to establish a new CLIA-certified diagnostic laboratory was also obtained from the Georgia DPH the week before student re-entry began on August 1^st^. Individual samples were retained for follow-up CLIA diagnostic testing of presumptive positives identified in initial surveillance; this two-step strategy reduced costs and increased volume.

A novel double-pooling scheme was designed to evaluate 15 individuals in six wells of a 96-well plate where each sample is present in two of the wells in a unique combination that, given prevalence around 1%, usually allows us to infer which sample is positive. This approach reduces false positives and controls the number of follow-up diagnostic tests, reducing costs. Leaving six wells for controls, 15×15 = 225 individuals are surveyed per plate, using Integra robots to generate pools, Kingfisher robots to isolate RNA, and QuantStudio quantitative RT-PCR testing. The actual PCR test was the CDC-approved IDEXX kit supplied by OptiMed, with control human RP and combined N1 and N2 SARS-CoV-2 primers. Individuals in negative pools are informed by email, usually within 24 hours, that “no further action is required” without giving an actual result, while presumptive positives, ambiguous samples, and some negatives are evaluated with CLIA-approved diagnostic tests. All positives were referred to Stamps Health Services (Georgia Tech’s health care center), reported to the Georgia DPH, and contact tracing was initiated. Students were given the option to return home or preferably to isolate for 10 days in a local hotel contracted for this purpose. All staff and faculty positives were reported to the Georgia DPH as required, contract tracing was initiated, and individuals were advised to consult a physician and isolate at home for 10 days.

In addition to the asymptomatic surveillance program, three other sources of tests were available for symptomatic individuals. Stamps Health Services provided Rapid LAMP tests for up to 20 students a day (with observed positivity between 10% and 20% across the semester), commercial Vault tests were available for some staff and on weekends, and some members of the Georgia Tech community preferred to be tested off campus with a variety of commercially available options. A Dashboard,https://health.gatech.edu/coronavirus/health-alerts visualizes daily case numbers by type of test or campus community, surveillance positivity, and tests per day.

## Results

A total of 1,825 positive tests were reported combining all testing modes. Figure 1 shows the number of cases per day (including both surveillance positives and total positives), along with a 7-day sliding average across the semester. These 1,825 positive tests corresponded to 1,508 individual cases since 236 participants tested positive twice, 31 thrice, and 6 four or more times. Most repeat positive tests were within a day or two of one another, however, multiple cases of positivity three or more months apart were observed. There is no indication that these are indicative of infectiousness, nor is there any information on whether they represent repeat infections. The vast majority of positive cases were students (1,351, 90% of cases, 9.7% cumulative positive) with just 157 Staff/Faculty infections of the 4,078 tested (3.8% cumulative positive), whereas approximately 15% of the tests were of Staff/Faculty. The surveillance program recorded 940 independent cases from 112,500 tests, an average positivity of 0.84%, and the cumulative estimated proportion of infected individuals was 8.4% based on approximately 18,029 registered participants. A large fraction of the campus community did not engage, either avoiding testing, or more commonly not visiting campus, namely working virtually and taking online instruction only.

**Figure 1.**
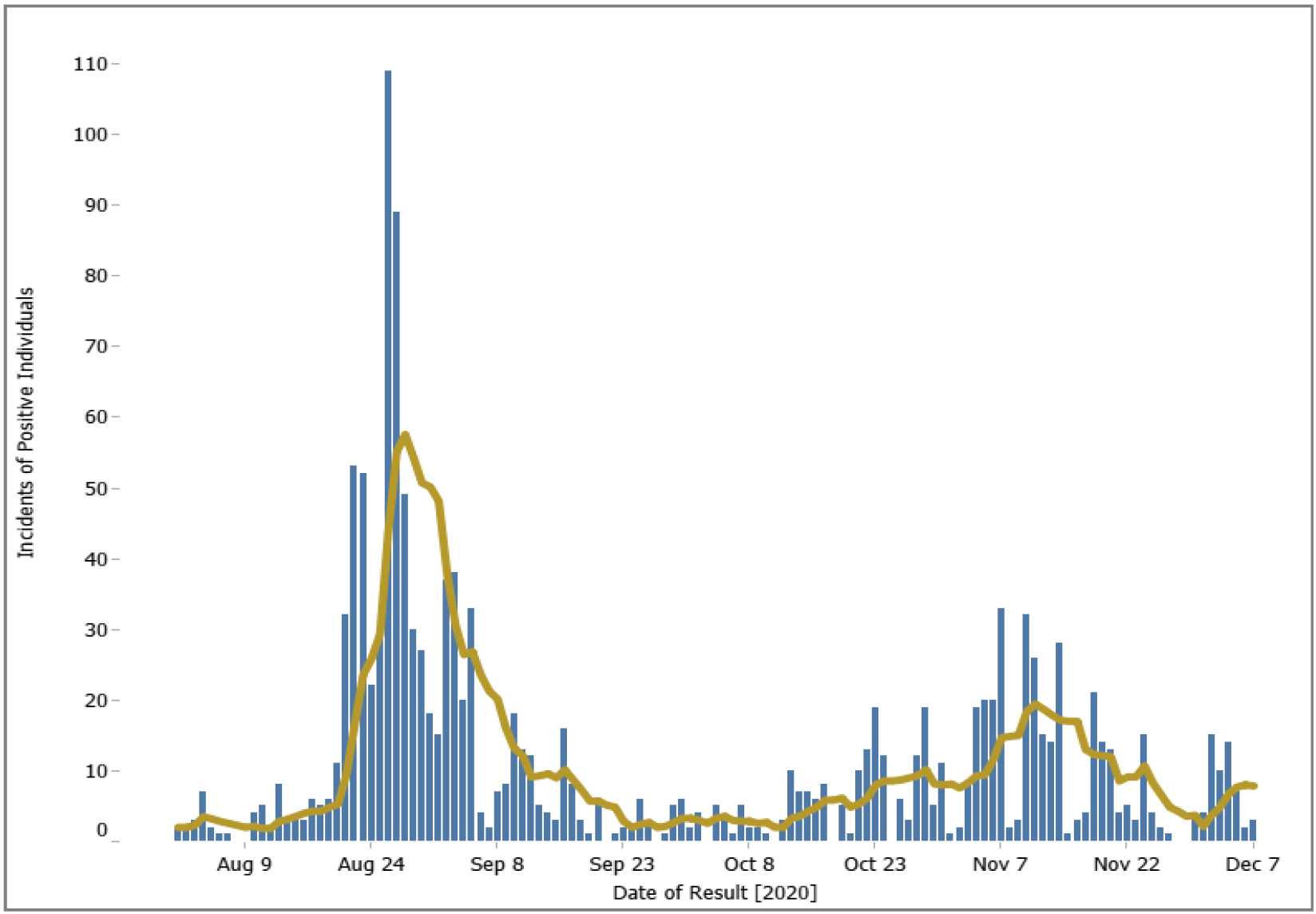
Number of positives from all tests per day (blue) with 7-day sliding window (yellow). Updates available at https://health.gatech.edu/coronavirus/health-alerts, which also allows interactive parsing of data by groups (residence, staff/student) and visualization of testing rates and positivity.

Three notable aspects of the results provide implications of public health relevance. The first is the ability of large-scale testing to rapidly identify and control case surges, as revealed twice in Fall 2020. After a low initial reentry positivity fraction under 0.5%, a severe outbreak occurred in week 3. Seven symptomatic cases were identified in one Greek house, which was followed by transmission through much of the Greek community and into nearby Residence Halls. An intensive campaign was initiated to test almost 90% of the on-campus residents over the next two weeks resulting in a peak asymptomatic positivity rate of 4%. Due, in part, to rapid case identification and isolation, the asymptomatic positivity rate steadily declined, and by mid-September positivity was consistently below 0.5% and on some days no positives were detected in more than 1,500 tests. Case rates increased in October, concordant with increasing community levels of transmission in Georgia, with asymptomatic positivity rate approaching 2% on a single day in late October. This second wave was more diffuse than the first and included a higher proportion of off-campus participants (likely due to community transmission unrelated to on-campus spread, noting that GT is located in Midtown Atlanta). Targeted surveillance testing of the most-affected dormitories was carried out in early November. Positivity declined to less than 0.5% in the final days of the Fall semester, again illustrating how focused testing can be part of an integrative strategy to reduce infections.

Our second observation is that there is heterogeneity with respect to Covid-19 awareness and risk. This observation is consistent with growing recognition of the importance of over-dispersion in transmission (7). The heterogeneity of risk is reflected in cumulative incidence data through September 25th summarized in Figure 2. Panel A illustrates that 75% of the positive tests were attributed to 25% of the Residence Halls or Greek houses, and reciprocally that 20%-33% of these living residences had almost no cases. Testing rates in Residence Halls was approximately 55% weekly after mid-Semester. Testing rates in Greek houses was approximately 75% weekly after mid-September, though it is notable that a subset of Greek houses had nearly 100% weekly participation whereas a few had <20% participation. The majority of student cases were restricted to the east side of campus in residences located proximal to the Greek community. Panel B contrasts the number of tests per individual through November 6^th^, for all individuals who had at least one test, showing clear bimodality in the general community (top), with a left-shift toward lower testing rates for the individuals who at some point had a positive test (bottom). Note that both distributions are subject to joint effects of sampling biases, confirmation bias (given that individuals with positive cases are recommended not to test again for a three month period), as well as intrinsic variation in individual risk and tendency to test. Nonetheless, we interpret both datasets as indicative of different behavioral approaches to the pandemic, with variation in the level of engagement with the testing program and practices that minimized individual risk. Although 44% of the individuals in Panel B tested at least biweekly (7 times or more), some groups of individuals refused to test at all or tested off-campus.

**Figure 2.**
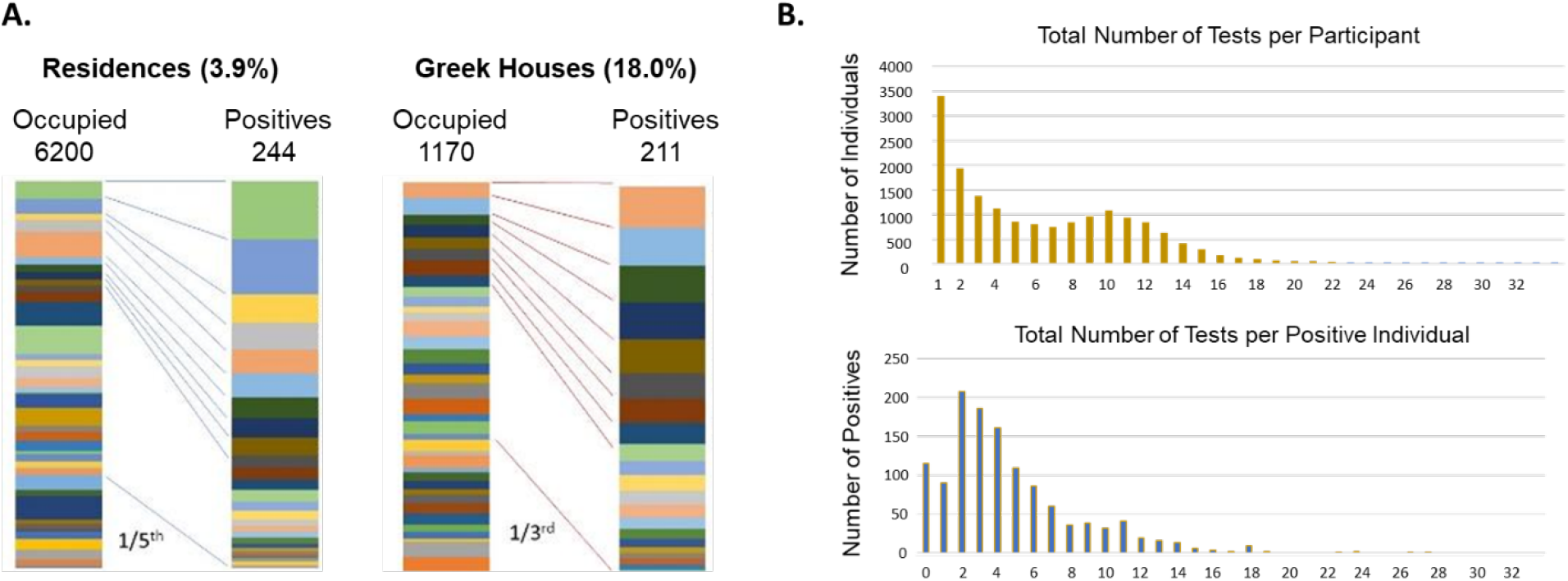
Positivity is heterogeneously distributed. (A) In both the Residence Halls and Greek houses by September 25th, almost 4% and 18% of individuals had been infected, but most of the cases were due to a few residences. In each pair of columns, the proportion of occupants or positives is represented by a different color for each residence. (B) Histogram distributions of the number of individuals testing (top) or diagnosed positive (bottom) who took from 1 to 32 tests across most of the Fall semester, August 9 through November 6.

A third observation was that shared double rooms had significantly elevated positivity. During the first peak, when one individual of a double tested positive, the second also tested positive over 30% of the time (38 of 125), generally within 3 to 5 days. Consequently, almost one half of the first wave cases were in doubles. There were several instances of a mini cluster arising in an adjacent block of rooms. Subsequently, all students were offered the option of moving to their own single, but many chose to remain with doubles, citing the preference to have more social interaction, and mental health concerns. Note that the finding of elevated infection rates in shared living spaces is consistent with findings of elevated household transmission [*8*].

## Discussion

Our results support the use of large-scale surveillance as a form of mitigation on college campuses, but contrast in important ways with other reports. Duke University for example reported just 84 total positives from a total of 68,913 tests performed on 10,265 students, only one third of which were detected directly from pooled surveillance, the remainder being discovered through contact tracing or as symptomatic cases (*9*). Their weekly positivity rate averaged just 0.08, though it should be noted that they report that just 18.4% of their positive pools led to a confirmed case, whereas our conversion rate for double-positive wells was very close to 100%. A survey of nine Boston-area Colleges (*10*) where testing of all resident students was performed at least weekly found a cumulative positivity of 1.6% (range 0.3% to 3.0%), also at least five-fold lower than the Georgia Tech rate. Note that documented new cases per capita in August were at least 3x higher in Atlanta than in Boston, though documented new case rates were more similar in September and October. The Boston study found no relationship between dormitory occupancy and positivity and no evidence for transmission within on-campus student housing (*10*), in direct contradiction to our conclusions.

The closest contextual comparison is provided by the University of Georgia (UGA), which reported (https://www.uhs.uga.edu/healthtopics/covid-19-health-and-exposure-updates) that the positivity rate exceeded 9% during their 3^rd^ week of testing and remained over 1% until mid-November, when another post-Thanksgiving increase to in excess of 4% occurred. In total, UGA reported 819 surveillance positives out of 33,435 tests for an average positivity rate of 2.5%. Given presumed similarity in NPIs, this contrast in outcomes provides some evidence for direct reduction of transmission via large-scale testing, namely a three-fold higher level of testing (at least five-fold relative to community size) and three-fold lower positivity fraction at GT relative to that of UGA. Notably, dashboard results from the University of Illinois Urbana Champaign (https://splunk-public.machinedata.illinois.edu/en-US/app/uofi_shield_public_APP/home) reveal more than 4,500 positive cases out of more than 1,000,000 saliva-based surveillance tests in the Fall semester, for an aggregate 0.44% positivity; that is approximately one-half of GT’s positive % with approximately 9-times as much surveillance testing. These contrasting examples provide context for efforts to assess costs and benefits in the implementation of large-scale viral surveillance testing.

In summary, focused testing of high-risk campus residences, including data-driven testing targeted at the level of individual floors and even clusters of adjacent rooms, along with NPIs and sustained communication were critical to GT’s ability to reduce transmission even amidst elevated community spread. Implementation of large-scale testing programs that include targeted increases of testing to respond to clustered outbreaks represent an opportunity to mitigate spread amidst college re-openings.

## Data Availability

No individual data is available since this was a surveillance study. Summary data can be downloaded from https://health.gatech.edu/coronavirus/health-alerts

## Acknowledgements

The authors are extremely grateful to the students and staff who embraced the program, and to dozens of colleagues in housing, communications, legal, and the PCR lab, who volunteered their time and, in many cases, redirected their effort to implement this program. Particular thanks go to Frank Neville, Aisha Oliver-Staley, Emily Ryan, True Merrill, and German Khunteev for their critical roles in establishing the program.

## Author contributions

GG, JSW, and MPS designed the program. MPS and MF performed the studies leading to EUA for the saliva test. MPS and AB ran the molecular testing lab. JAW and AO supervised sampling and logistics. BL and SB were responsible for data handling. BH was the medical director. CTA and AJG supervised the program and provided administrative support. GG and JSW wrote the paper.

